# Efficient sample pooling strategies for COVID-19 data gathering

**DOI:** 10.1101/2020.04.05.20054445

**Authors:** Istvan Szapudi

## Abstract

Sample pooling of CoViD-19 PCR tests has been recently proposed as a low cost alternative to individual tests. We show that sample pooling is efficient as long as the fraction of the population infected is relatively small. Fisher information theory suggests a rule of thumb that for low infection rates *p*, pooling 2*/p* samples is close to optimal. We present a simple strategy for survey design when not even a ballpark estimate of the infection rate is available.

## Introduction

As the CoViD-19 pandemic sweeps through the planet, data gathering is desperately needed for understanding the spread of the disease, the fraction of the population infected, and the case fatality rate (CFR) and infection fatality rate (IFR) for each age group (1–3). In most countries, only symptomatic individuals are tested referred by their attending physicians due to the scarcity of tests.The bottleneck is the expense and availability of the PCR test kits for the SARS-CoV-2 virus. Yet, massive surveys could make a big impact on decisions about influencing the behavior of the populace to mitigate the effects of the pandemic in a non-pharmaceutical fashion, such as invoking a general or partial lock down.

Sample pooling, carried out as mixing of samples, e.g., saliva, from several individuals, provides a cost effective solution for massive testing of the population at large. Recently, a protocol has been developed for sample pooling for CoViD-19 tests (4, 5). In addition, a group proposes to test the whole population of Hungary in batches of 15-64(6). Sample pooling has been considered since World War II. for a wide variety of applications, (e.g., 7, 8, and references therein). In these notes we perform the fundamental calculations to support the design of efficient surveys with pooled samples.

### The Fisher information of pooled sampling

We introduce a simple model to quantify the aim of a CoViD-19 survey: in a particular age bracket, let’s assume that a fraction *p* of the population is infected. We want to measure this fraction. The fraction of the population not infected is *q* = 1 − *p*. Let us assume that in our survey we pool the samples of *n* persons. For such a test to be negative, the probability is *P*_−_ = *q*^*n*^, and consequently, the probability that the test yields a positive result is *P*_+_ = 1 − *q*^*n*^. As a consequence, after *N* measurements, the probability of finding *N*_+_ positive and *N*_−_ results is

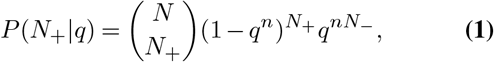

where *N* = *N*_+_ + *N*_−_ by definition. This formula assumes the independence of each test from each, and neglects any correlations between each pool (e.g. when families are tested in one pool, there results are more likely to be positive or negative together). For intuitive picture, the formula is equivalent to coin tossing, assuming the probability of heads is *q*^*n*^.

We wish to use Bayesian inference to extract the information the data has on *q* (and therefore *p* = 1 − *q*). Using Bayes’ theorem, the likelihood of *q* is

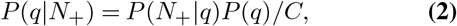

where *C* is normalization of the probability distribution. We can use a non-informative prior on q between [0, 1], and absorb that constant into *C*. The above function can be used for numerical analysis of data. If several different pools have been used, the likelihood functions simply multiply. Maximizing the log likelihood will yield the most likely value of *q*, and the confidence intervals can be determined the standard way.

Some analytical results can be obtained as well. The most likely value of *q* can be found taking the derivative of the log likelihood

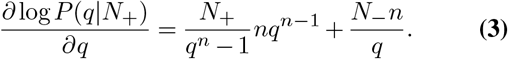

Equating the above equation with zero yields *q*^*n*^ = *N*_−_*/N* is expected. The second derivative of the log likelihood is the curvature

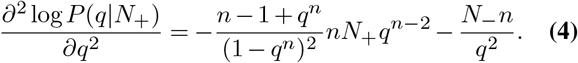

Taking the negative expectation value yields the Fisher information matrix:

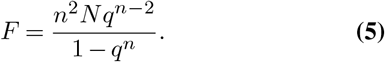

The variance of the measurement is 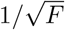. For *n* = 1 we recover the standard result of *F* = *N/q*(1 − *q*).

### Optimization

For a given amount of resources allowing *N* tests total, the Fisher information can be maximized for *n* if we have a preconception about q. Taking the derivative of the previous equation according to *n*

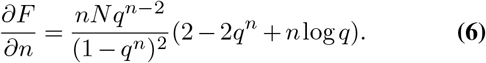

Equating the above with zero, the equation in the parenthesis has to be solved numerically. Since it depends only on *q*^*n*^, a universal solution is *q*^*n*^ = 0.203188. Table 1 contains the values of q for practical values of sampling pooling for CoViD-19.

**Table 1.** Optimal batch size for pooling as a function of *q* = 1 − *p* expressed as a percentage for convenience.

| $n$ | $q$ |
| --- | --- |
| 64 | 97.5% |
| 32 | 95% |
| 16 | 90% |
| 8 | 82% |
| 4 | 67% |
| 2 | 45% |

## Conclusions and Discussion

To measure the infection rate in a particular age bracket, samples should be pooled based on the results of Table 1. For final results, the a full maximum likelihood analysis of the data is recommended.

If there is no reasonable estimate of the infection rate is available to guide the design of the pooled survey, one case start with large pool, i.e. *n* = 64. If after *N* tests, all measurements are positive, it means that the infection rate is too large for this pool. If *N* = *N*_+_, the most likely value for *q* = 0, therefore Eq. 5 cannot be used to estimate confidence intervals. In that case the likelihood function is (1 − *q*^*n*^)*N*, and one has to integrate it directly to obtain a confidence interval for *q*. This can be done numerically, as an example, assuming that 10 measurements produced positive results with *n* = 64 sampling, there is a 94% chance that *q ≤* 0.9, i.e. one should try *n* = 16 next. Thus a simple strategy would be to determine the optimal pooling rate with preliminary measurements of higher pooling rate than necessary, and design the survey with the closest rate found in Table 1. Note also that when the rates are low, individuals can be pinpointed with a binary search with log *n* extra measurements, but when the infection rate is high, individual measurements are more efficient (5).

Note that realistic protocols realising the idea will have false positives and negatives. These should be calibrated and taken into account when an actual measurement is translated into IFRs. These issues are straightforward fold into the likelihood function in a forward Bayesian analysis, as proposed here. On the other hand, the conclusions are robust enough to be useful for survey design.

Note that pooled sampling has other applications, such as testing for infection on a regular schedule in a group of people, e.g., health care workers on the same shift.

## Data Availability

All codes and calculations are available upon request.

## ACKNOWLEDGEMENTS

IS thanks Istvan Csabai and Carlo Graziani for useful comments.

